# Monitoring the evolution of the COVID-19 pandemic in China, South Korea, Italy and USA through the net relative rate of infection of the total number of confirmed cases

**DOI:** 10.1101/2020.05.15.20103259

**Authors:** João Manoel Losada Moreira

## Abstract

Managing the COVID-19 pandemic in the middle of the events requires real-time monitoring of its evolution to perform analyses of containment actions and to project near future scenarios. This work proposes a scheme to monitor the temporal evolution of the COVID-19 pandemic using the time series of its total number of confirmed cases in a given region. The monitored parameter is the spread rate obtained from this time series (day^−1^) expressed in %/day. The scheme’s capability is verified using the epidemic data from China and South Korea. Its projection capability is shown for Italy and United States with scenarios for the ensuing 30 days from April 2^nd^, 2020. The spread rate (relative rate of change of the time series) is very sensitive to sudden changes in the epidemic evolution and can be used to monitor in real-time the effectiveness of containment actions. The logarithm of this variable allows identifying clear trends of the evolution of the COVID-10 epidemic in these countries. The spread rate calculated from the number of confirmed cases of infection is interpreted as a probability per unit of time of virus infection and containment actions. Its product with the number of confirmed cases of infections yields the number of new cases per day. The stabilization and control of the epidemic for China and South Korea appear to occur for values of this parameter below 0·77 %/day (doubling time of 90 days).

## Introduction

The COVID-19 exponential pandemic has spread across the world, affecting society, health care systems and governments in different manners.^1–4^ Studies about monitoring the dynamics of the COVID-19 pandemic in China and other countries are found in the literature.^5–9^ Most of them assess the evolution of the pandemic transmission process through its reproduction number, R, inferred in different moment, and correlate variations of the results with interventions conducted to contain its spread. The decline of the reproduction number is associated to intervention actions.^5–9^ Studies focusing more directly on interventions to contain the pandemic performed simulations based on the SIR and SEIR models and make assumptions about parameters of interaction between the population compartments of susceptible, exposed, infected and recovered individuals.^10–13^ These studies allow understanding the pandemic evolution in possible scenarios but does not tackle directly the problem of real-time monitoring.

Other studies try aim at approaches to account for spatial and temporal effects of the pandemic evolution.^14–16^ Despite the difficulty to perform real-time monitoring Chen and Yu^15^ propose a scheme for regions in China based on extracting information from the second derivative of the time series of the total number of infections which they interpreted as an indicator of the acceleration or deceleration of the infection process. They report success in identifying the impact of containment measures on the infection spread.^15^ For temporal monitoring purposes, Park et al^16^ suggest the use of the spread rate (relative rate of change) of the time series of the incidence instead of the reproduction number which, as a unitless quantity, does not contain any information about time.

This work proposes a scheme to monitor the exponential evolution of the pandemic through the spread rate considering Park et al^16^ suggestion which is coincident with the industry practice with processes of similar dynamics or kinetics.^17–22^ The approach is to monitor the exponent of a key variable of the dynamic process under study.^29,20^ We take the time series of the number of confirmed cases of infection with the new coronavirus as the variable that can provide information about the dynamic behaviour of the pandemic. We interpret the results using a scheme based on the SIR model.^23^ The goal is to monitor the dynamic of the pandemic in real-time and being able to construct scenarios for assessing its evolution in the following 30 days.

This article begins presenting the data, methods and the theoretical basis for interpreting the results. Following, we present the results and discussions divided in two parts: in the first, we simulate the successful control of the pandemic carried out by China and South Korea attempting to verify how well the proposed scheme monitors these two pandemic processes. In the second part, we apply the scheme to Italy and United States, and present projection results of scenarios aiming at controlling the pandemic in these countries. Finally we present the conclusions.

## Data and methods

The COVID-19 epidemic spreads across a region and is detected through tests applied to the population in varied conditions.^9,10,13^ The relationship between the total number or population of infected individuals in a region, I_n_, at day t_n_, and the number of infected individuals counted or detected by the tests applied to the population, C_n_, is

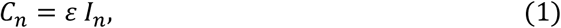

where ε is the overall sampling scale factor of the infection detection system (counts of confirmed cases in the region/true number of infected individuals in the region) and the sample has to be representative of the true infected population.^19–21^ Additionally, ratios such as lethality rate based on monitored confirmed cases can provide good estimates of hospital demands and other important time varying information. In this work we take the relative rate of change or exponential growth rate of C_n_, to model the pandemic temporal evolution because it is the time series with more data available in the early phase of the epidemic.

The data were obtained from the health systems of China, South Korea, Italy, and the United States which are organized as time series in the Worldometer site.^4^ Figure 1 shows the number of confirmed cases, C_n_, of COVID-19 for China, South Korea, Italy, and United States for the period from January 20^th^ to April 15^th^, 2020. In the abscissa day 0 is 2020/01/31. For China and South Korea it is possible to observe the flattening of the confirmed cases curve over time, with the number of cases in China being approximately an order of magnitude higher than the South Korean in late March. For Italy and US, the infection process is already showing signs of reduction and the same curve starts to flatten but the evolution in the number of cases is still unknown.

**Figure 1.**
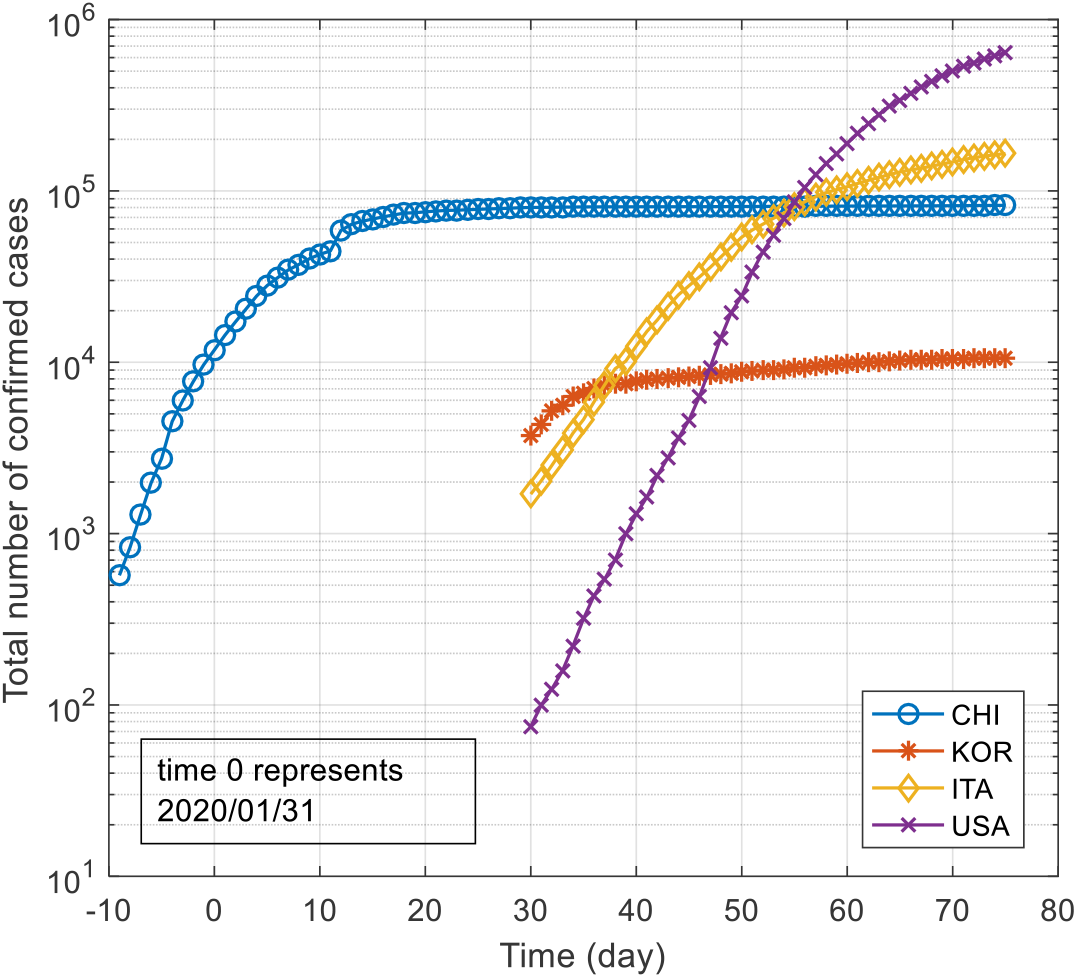
Total number of COVID-19 cases detected over time (C_n_) for China, South Korea, Italy, and United States.^4^

The problem posed is obtaining from these data dynamic information about the COVID-19 pandemic which is useful for those responsible to control it. The method adopted in this work follows the steps below:

a. Construct a model to infer the temporal evolution of the pandemic using the relative rate of change over time, *a_n_*, obtained from C_n_;
b. Establish scenarios for the temporal evolution of the number of new cases of infection (NC_n_) in the subsequent 30 days.

In the rest of the article we use the words pandemic and epidemic interchangeably.

### Temporal model based on the relative rate of change of the number of cases of COVID-19

The balance equation for number of infected individuals according to the SIR model^9,13,23^ is

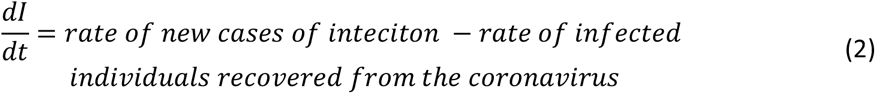

The rate of new cases of infection depends on the action of the virus and the containment measures taken by society to avoid the spreading such as different of social isolation, quarantine, hygienic measures etc. This infection rate, *a_i_*(*t*), is assumed to be proportional to the number of infected individuals on day t, I(t), who act as spreading agents of the infection. We can define *a_i_*(*t*) as a probability of new infection per unit time and per infected individual to determine the infection rate.

The rate of removal of infected individuals depends on recovery from the infection and death occurrences. If we assume that this removal rate, *a_r_*(*t*) is also proportional to the number of infected individuals, we can also define a probability of removal of infected individuals per unit time and per infected individual that takes into consideration all these removal processes. If we put together these two effects (new infection and removal) we obtain a probability of the net result of infection and removal per unit time and infected individual.^21,22^ Thus Eq. 2 can be written as

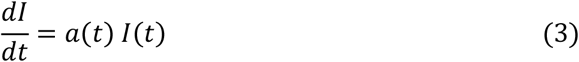

where *a*(*t*) = *a_i_*(*t*) − *a_r_*(*t*) is the probability of net infection and removal of infected individuals accounting for new infection of individuals, containment actions and removal of infected individuals from society. The net rate of infections appearing in the region, *NC*(*t*), would be equal to the product *a*(*t*)*I*(*t*).

Taking the time interval of one day, the solution of Eq. 3 is given by an exponential evolution of the number of infected individuals, that is,

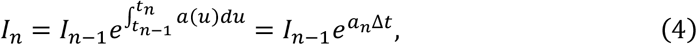

where t_n_ is the n_th_ day, *a_n_* is the average value of a(t) on the n_th_ day, and I_n_ is the number of confirmed cases on day t_n_, and Δt = t_n_-t_n-1_ or one day.

*a_n_* is the spread rate of transmission of the epidemic, because when its value is positive I_n_ grows exponentially, when its value is negative, I_n_ falls exponentially and when it is null I_n_ is constant over time.^12^ *a_n_* can be estimated at t_n_ by

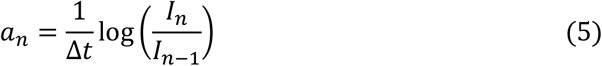

This probability of net infection per unit of time and per infected individual would have to be obtained from a time series of the active number of infected individuals. Since this time series is not accurate in the middle of the epidemic process, we approximate the time variation of the active number of infected individuals as that of C_n_. In addition, we assume that the scale factor, ε, is invariant over time and adequately covers the region affected by the epidemic. Thus we approximate *a_n_* as

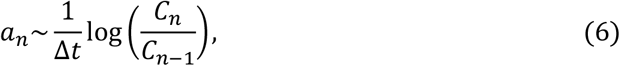

and the number of new cases per day at t_n_ as

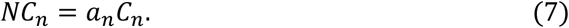

It should be noted that *a_n_* given by Eq. 6 is nothing more than the net relative rate of change in the total number of confirmed cases of infection calculated considering continuous time variation^11–13^ It is a partial net relative rate of infection because it accounts for the transmission action of the virus and measures to contain it such as social isolation or cleaning hands taken by the individuals. It does not account for those individuals who were infected and recovered from the illness.

The approximations of Eqs. 6 and 7 produce interesting consequences. First, since *C_n_* is a cumulative variable it always grows. Thus *a_n_* is always positive and the steady state condition for Eq. 3 is obtained only when *a_n_* = 0. Second, due to Eq. 7, we have that *a_n_* multiplied by *C_n_* yields the number of new cases *NC_n_* at time *t_n_*.

In the rest of this article we refer to *a_n_* interchangeably as the net probability of infection per unit of time or net relative rate of infection and express it in day^−1^ or %/day. The doubling time of infection, td_n_, is related to the net rate of infection as 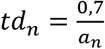. To reduce the statistical variation of the data, a 3-day moving average was applied to the data before obtaining the net relative infection rates.

The main approximations of this monitoring scheme are:^21,22^ a) taking C_n_ as the active number of infected cases which is valid during the first months of the pandemic; and b) considering the scale factor, ε, constant throughout the study. Regarding the first approximation, it is a good approximation in the beginning of the epidemic when its growth is exponential and much bigger than the recovery rate. Regarding the second approximation, if it is not verified the analyst must consider the results with care.

## Results

### Monitoring of coronavirus containment actions in China and South Korea using the net relative infection rate

Figure 2 shows the net relative infection rate for China and South Korea obtained through Eq. 6 using the data presented in Figure 1. The x-axis shows the days and the day 0 corresponds to 2020/01/31. The y-axis is presented on a logarithmic scale to ease comparing the rates of change in China and South Korea along the studied period. The straight line represents a condition of net relative infection rate of 0·77 %/day or 90 days of doubling time, a period sufficiently long to control it and plan and take actions to provide needed infrastructure to national health systems. In other words an epidemic condition in which the health systems can control it and manage its consequences.

Figure 2 shows that in China, before actions of social containment in January 2020, the net probability of infection or net relative rate of infection was 40 %/day. After the containment actions started it dropped to between 0·04 and 0·1 %day. South Korea started rapidly its containment action (beginning of March) and its net relative rate of infection dropped and stabilized around 1 %/day and then around 0·3 %/day. The epidemic condition of this two countries was one of very slow transmission rate (very low net relative rate of infection) with doubling times between 200 and 2000 days.

**Figure 2.**
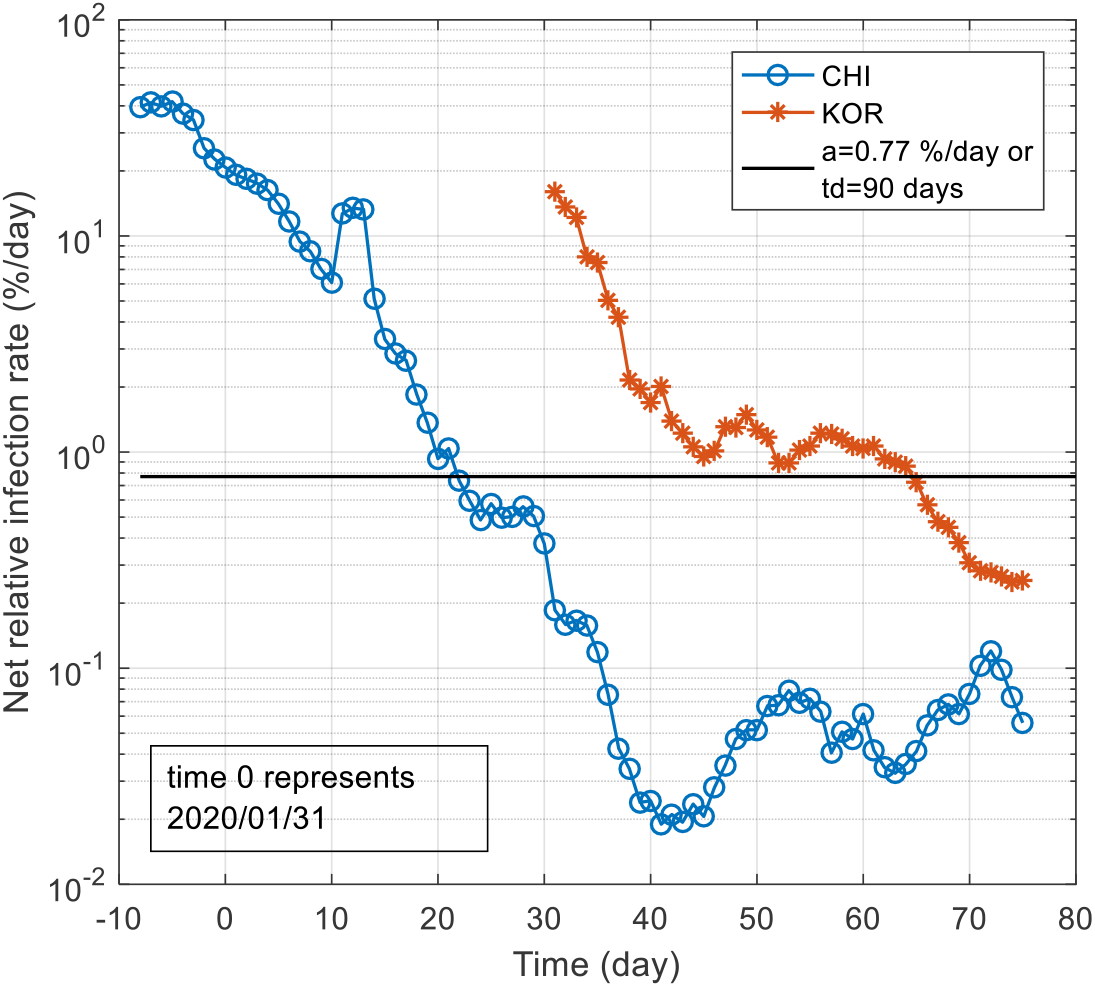
Net relative infection rate as a function of time for China and South Korea.

### Estimate of new cases of infection per day over time

Figure 3 compares the number of new cases per day estimated by Eq. 7 with the new cases obtained from the reported data. NC_n_, given by Eq. 7, reproduces the overall behaviour of the true number of new confirmed cases per day. The root mean square of the relative discrepancy for China was 37 % and for South Korea, 51 %.

Figure 3 shows that after day 65 (mid-April), when the *a_n_* for both countries were below the 90 day doubling time line (Figure 2), the number of new cases in China and South Korea oscillated around 60.

**Figure 3.**
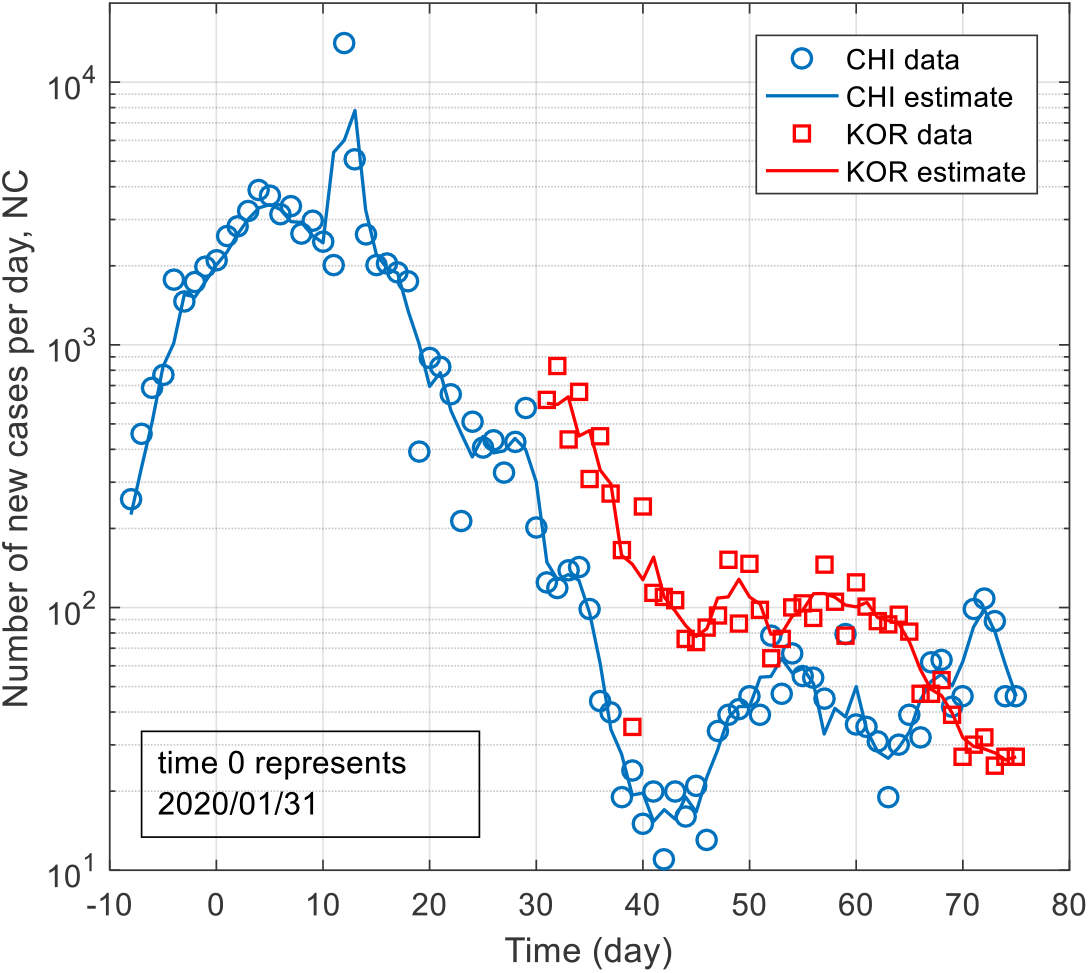
Comparison between the number of detected new cases reported in the data and the rate of detected new cases estimated by Eq. 3.

### Epidemic monitoring and projections for Italy and the USA

Figure 4 shows the net relative rate of infection by the coronavirus for Italy, the United States, and South Korea taken now as a successful benchmark process to control the epidemic. The curves for Italy and USA show similar behaviour when compared to China. They started with net relative rates of infection between 20 and 30 %/day and after introducing measures of social isolation their figures began to drop.

**Figure 4.**
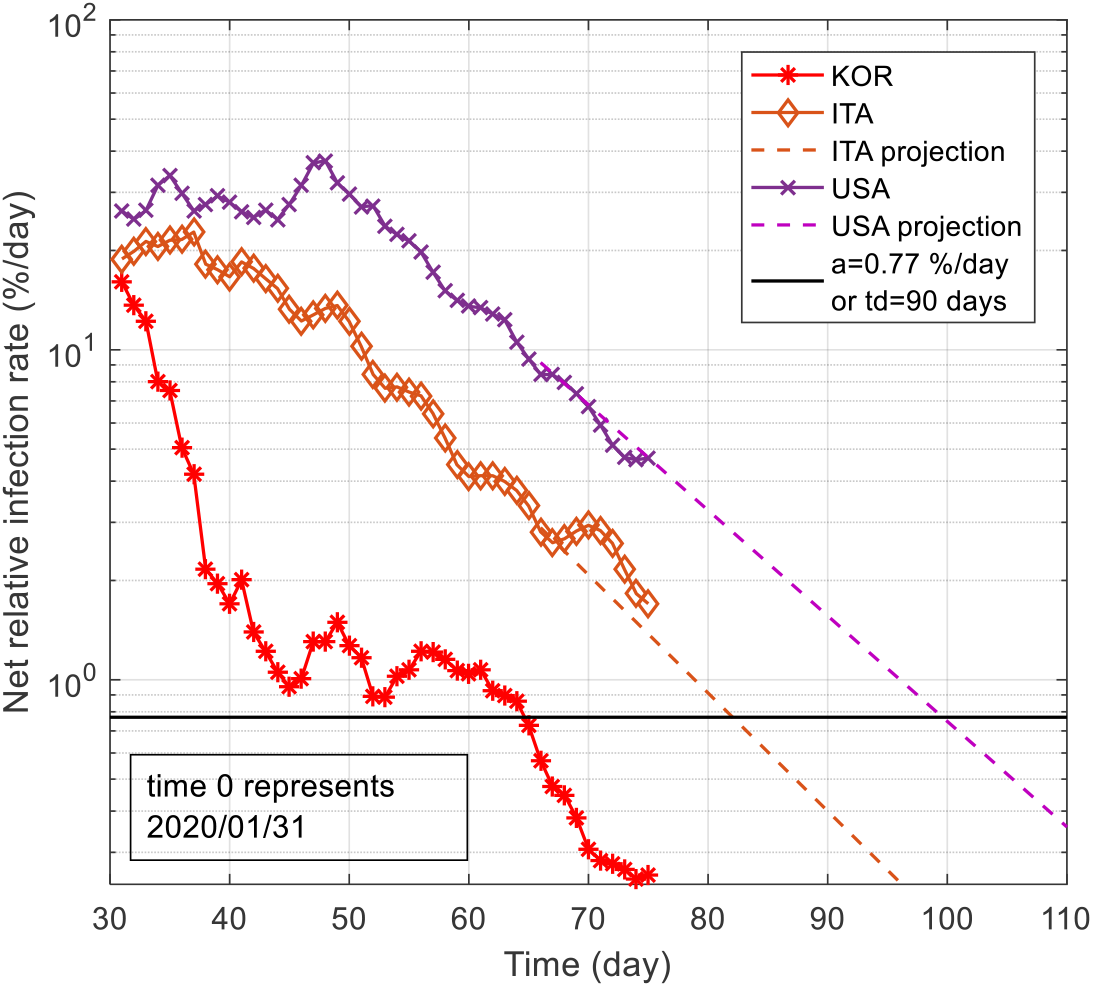
Net relative infection rate as a function of time for South Korea, Italy and USA and linear projected values for Italy and USA. The projections were performed based on data from day 50 to day 63.

The figure also presents projections for Italy and the United States based on the linear extrapolation of the previous behaviour between the days 50 and 62, that is, between March 21^st^ and April 2^nd^, 2020. The slopes of the extrapolations are –0·0856 %/day^2^ for Italy and –0·0730 %/day^2^ for the USA. The linear extrapolation of the net relative rate of infection of these two countries, considering that their respective behaviours in these 13 days are repeated, indicates that Italy and the United States cross the 90 day doubling time line around the days 80 and 100, respectively. It should be noted that such results represent a scenario that may occur only if the previous conditions persist.

The figure shows the projected estimations for C_n_ and NC_n_ for the next 30 days. The projected values for C_n_ and NC_n_ can be compared to actual data between days 62 and 75, or April 3^rd^ and April 15^th^.

The projected net relative rates of infection allow estimation of future C_n_ and NC_n_ through Eqs. 4 and 7, and Figures 5 and 6 present these results for Italy and China, respectively. When Italy reaches the 0·77 %/day line on day 80 its figures for these two variables would be around 200,000 total cases and 1000 new cases per day, respectively. For the USA, on the day 100, the same figures would be around 1 million and 10,000, respectively.

**Figure 5.**
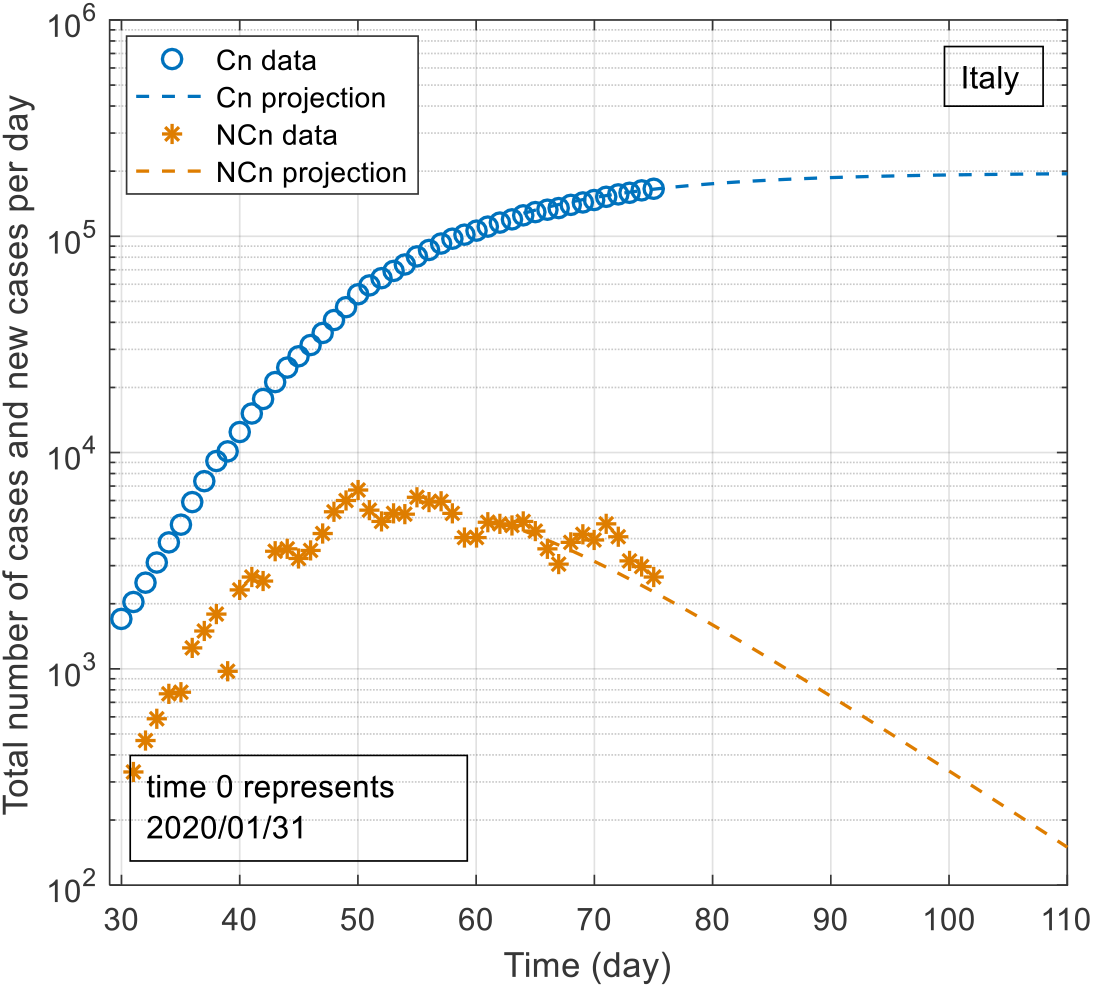
Projections of the total number of confirmed cases and the number of new cases per day for Italy. The projections, in dashed lines, were calculated using Eqs. 4 and 7.

**Figure 6.**
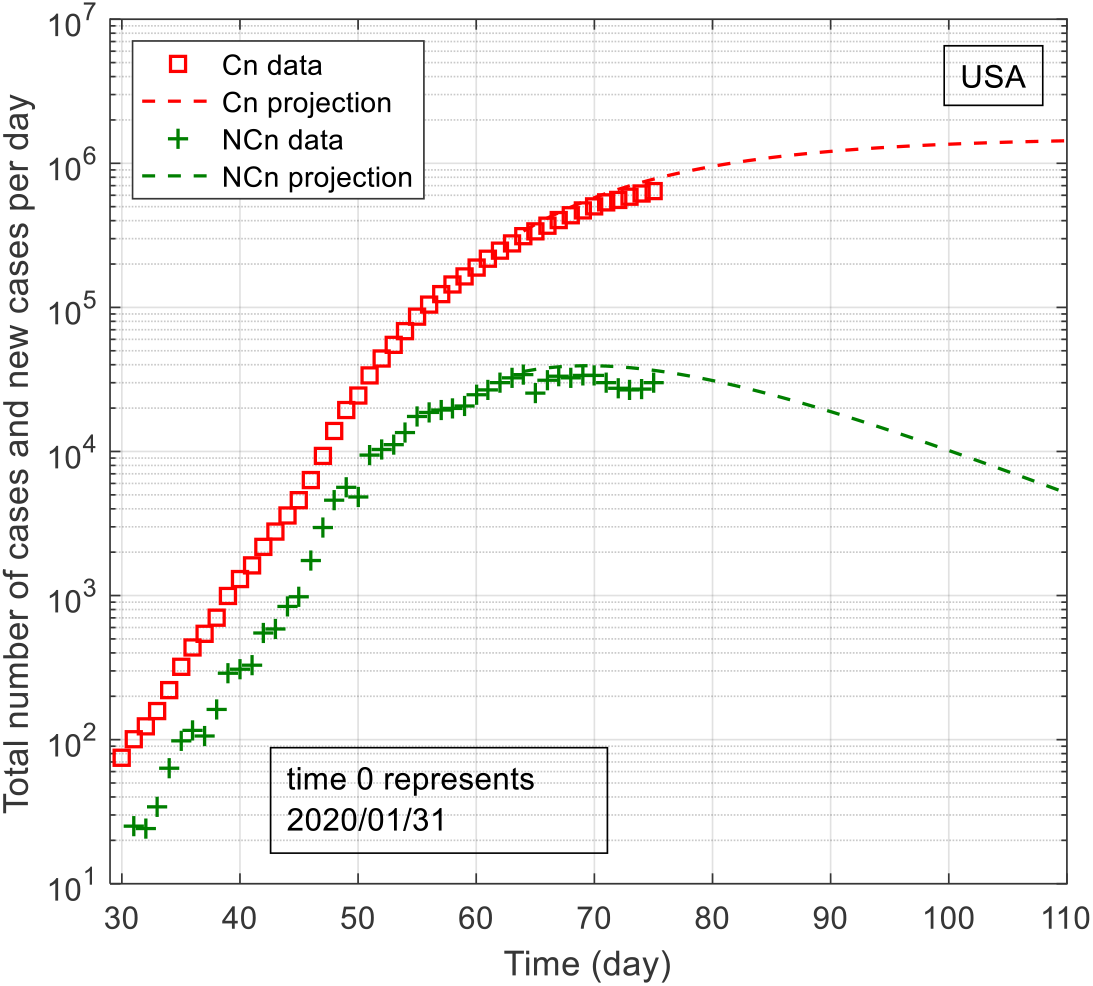
Projections of the total number of confirmed cases and the number of new cases per day for the USA. The projections, in dashed lines, were calculated using Eqs. 4 and 7.

## Discussion

### Capability of the proposed scheme to monitor the coronavirus containment actions in China and South Korea

The net relative rates of infection for China and South Korea shown in Figure 2 allow us to monitor the effectiveness of measures to contain the spread of the virus in this two countries, i.e., how fast the relative rate of infection falls. In China, the net relative rate of infection was 40 %/day and that country started its measures to control the spread of infection in their society. At this time, the doubling time of the number of infected persons was between 2 and 3 days. The measures of social isolation were very strong and produced a continuous reduction of a_n_. On February 25^th^, the rate was already less than 1 %/day (70 days of doubling time) and on March 8, less than 0·1 %/day (700 days of doubling time or about 2 years). On 03/27/2020 the confirmed number of infected people in China was 81394, the net relative infection rate was 0·06 %/day and the number of new cases per day, very small.

The infection reached the other countries strongly in March 2020. In South Korea, the infection process started a little earlier and the country took drastic measures to contain the spread of the virus, including social isolation. In early March, South Korea’s net relative infection rate was just below 20 %/day (3·5 days of doubling time). After March 15^th^ it stabilized around 1 %/day (70 days of doubling time) and on 03/27/2020 the confirmed number of infected persons was 9332. Why did this country decide to stabilize initially its infection rate around 1 %/day and not around 0·05 %/day as China did?

The answer to this question is the number of new cases per day, NC_n_, in both countries which were between 50 and 100 and could be directly correlated to hospital demands. These figures seemed suitable to handle the consequences of the epidemic on their respective society.

The important variable is NC_n_, the rate of new confirmed cases per day, obtained from the product *a_n_C_n_*, Eq. 7. Given that the total number of infected cases for China was around 10 times greater than that of South Korea, it need to lower its net relative rate of infection to levels 10 times lower to obtain similar numbers of new cases. It actually dropped more than that (see Figure 2).

More importantly, Figure 2 shows that the logarithm of the net relative rate of infection is very sensitive to sudden changes of the epidemic evolution and thus it can be used to monitor the effectiveness of containment actions of the epidemic. Additionally, it allows one to identify clear trends of the epidemic process and project future scenarios for its evolution.

### Epidemic monitoring and projections for Italy and the USA

Figure 4 allows comparison of the evolution of the epidemic in Italy and USA with that of South Korea. The figure shows clearly that South Korea managed to contain the epidemic more rapidly than Italy and USA. It shows also that the signs of reduction of the epidemic in South Korea evidenced about 10 days before than in Italy and 15 days before than in the USA. Given the high net relative rates of infection (around 20 %/day) of Italy and USA, these delays caused important increases in their number of confirmed infected cases.

The results of Figures 2 and 4 show that the logarithmic display ease the identification of the course of the epidemic. While the number of confirmed cases and new cases per day are increasing (see Figure 1), these figures show clearly if the epidemic is being controlled or not.

### Accuracy and utility of the proposed monitoring scheme

Regarding Figure 3, the relative discrepancies of 37 % and 51 % for China and South Korea data can be explained mostly by the scattering of the reported data of new cases per day. Comparing the results with 3-day average of the new cases per day data would have presented much better comparison. Figure 3 shows clearly that the estimation of NC_n_ for both China and South Korea suffices for understanding the dynamic or kinetics of the epidemic and for decision taking.

In Figures 5 and 6, the difference between the linear extrapolation and the actual reported data between days 63 and 75 shows typical pointwise discrepancies one may face in doing such exercises. They show that the accuracy of the scheme is sufficient to identify evolution trends of the epidemic and produces meaningful estimates for projected number of new cases of infection per day. The results show that the total number of infected cases in Italy will stabilize around 200,000 and in USA around 1,000,000.

## Conclusions

The proposed scheme for monitoring the COVID-19 epidemic based on the net relative rate of change of the number of confirmed cases of COVID-19 infection presented good results. This parameter is very sensitive to sudden changes in the epidemic evolution and a plot of is logarithm as a function of time provides a good visual picture of the epidemic dynamic evolution over time. The effects of containment measures are clearly identified but it is not possible to identify which one influenced the infection dynamics. Additionally, it presents clear trends of the epidemic process which can be used to project future scenarios for its evolution.

The proposed scheme is not a model that simulates the epidemic but a monitoring scheme that infer the current net relative rate of infection based on reported confirmed cases of infection. Scenarios can be constructed by assuming future trends for net relative rate of infection.

The stabilization and control of the epidemic appears to occur for values of the net relative rate of infection below 0·77 %/day or 90 days of doubling time of the total number of confirmed cases. The specific value, however, depends on the capacity of each country or region to take care of its fellow citizens and the confirmed number of persons infected. If the latter grows significantly, the target lower net relative rate of infection has to be lower and, consequently, the effort to contain the spread of the epidemic is greater. South Korea stabilized its infection process in ∼ 15 days and China in ∼ 50 days.

The main approximations of this monitoring scheme, which will be taken as future studies, are taking the total number of confirmed cases as the active number of infected cases, which is valid during the first months of the pandemic; and considering the scale factor, relating the true number of infected individuals to the detected number of confirmed cases in a region, invariant throughout the evaluation.

## Data Availability

All data are referred to in the manuscript. The data were collected from: https://www.worldometers.info/coronavirus/#countries

## Acknowledgments

The author thanks the Brazilian agency Coordenação de Aperfeiçoamento de Pessoal de Nível Superior (CAPES) for financial support and acknowledges it had no interference on data collection, interpretation, or decision to submit the work for publication.

